# The ARumenamide AR-788 suppresses the transient outward current in human iPSC-derived right-ventricular Brugada cardioids

**DOI:** 10.64898/2026.10.06.26364857

**Authors:** Yohan Guéret, Aurore Girardeau, Sarah Godin, Maureen Choteau, Jérôme Montnach, Guillaume Lamirault, Nathalie Gaborit, Mena Abdelsayed

## Abstract

Brugada syndrome (BrS) is an inherited arrhythmia disorder with limited drug therapy, quinidine being the principal agent. Loss-of-function variants in *SCN5A* reduce the cardiac sodium current (INa), but the arrhythmogenic imbalance also depends on the transient outward potassium current (Ito). Here we generated human induced pluripotent stem cell-derived cardioids with right-ventricular identity from a patient carrying an *SCN5A* frameshift variant (p.A665Gfs*16) and characterised them by patch-clamp and optical mapping. BrS cardioids showed 60% lower peak INa, more than doubled Ito, 38% slower conduction and 76% longer spontaneous cycle length. We then tested AR-788, a benzylaminothiazole– benzimidazole carboxamide of the ARumenamide class. In wild-type (WT) cardiomyocytes AR-788 inhibited Ito concentration-dependently (12%, 30% and 42% at 5, 10 and 20 µM; extrapolated IC50 25.4 µM), and at 10 µM it returned BrS Ito to the level of untreated WT cells (21.71 vs 21.48 pA/pF, p = 0.9716) without reducing INa. It also reduced the rapid delayed-rectifier current (IKr) tail current by 40%, a liability requiring formal proarrhythmia profiling. These data identify AR-788 as an Ito-suppressing member of the ARumenamide class and provide proof of concept that a small molecule can correct the potassium-current excess in a human BrS model.

## Introduction

Brugada syndrome (BrS) is a heritable primary electrical disease of the heart characterized by coved ST-segment elevation in the right precordial leads and a predisposition to polymorphic ventricular tachyarrhythmia and sudden cardiac death, typically in structurally normal hearts^1^. Loss-of-function variants in *SCN5A*, which encodes the pore-forming α-subunit of the cardiac voltage-gated sodium channel Nav1.5, are the most frequently identified genetic cause^2^. Despite decades of mechanistic work, therapeutic options remain narrow: implantable cardioverter-defibrillators address the consequences rather than the substrate^3^, and quinidine, valued largely for its inhibition of the transient outward potassium current (I_to_), remains the principal pharmacological option, constrained by tolerability and availability^4^.

Two mechanistic frameworks, the repolarization and depolarization hypotheses, converge on a common theme: a regionally accentuated imbalance between inward (depolarizing) and outward (repolarizing) currents during the early phases of the action potential^5,6^. A reduced I_Na_, combined with a prominent I_to_ in the right-ventricular epicardium, exaggerates the phase-1 notch, can precipitate loss of the action-potential dome, and creates the dispersion of repolarization that underlies phase-2 reentry^6,7^. This model places I_to_, not only I_Na_, at the center of the arrhythmogenic substrate and rationalizes why an agent that attenuates Ito can be antiarrhythmic even when the primary lesion is a sodium-channel deficit^4,7^. The model is chamber-specific: the substrate is a property of the right-ventricular outflow tract, where the epicardial I_to_ is largest.

Human induced pluripotent stem cell (hiPSC) models have made it possible to study these currents in a patient-specific, human cellular context^8,9^. Patient-derived cardiomyocytes reproduce the reduced sodium current and afterdepolarizations of BrS^9,10^, and three-dimensional cardioids further recapitulate multicellular electrical behavior, conduction and spontaneous rhythmicity, that monolayers capture only partially^11,12^, enabling both single-cell voltage-clamp and tissue-level optical mapping in the same model system. Because chamber identity can now be directed during differentiation, cardioids of right-ventricular character can be generated13,allowing the Brugada substrate to be modelled in the chamber in which it arises.

AR-788 (AR) belongs to the ARumenamide class of benzylaminothiazole carboxamides originally identified through structure-guided screening as modulators of Nav1.5, with the therapeutic hypothesis that they stabilize channel availability in loss-of-function settings14. The class is not, however, Nav1.5-selective: its best-characterised member, ARumenamide-787, was subsequently shown in canine ventricular myocytes and arterially perfused wedge preparations to inhibit Ito as its predominant action while enhancing INa15. AR-788 is a distinct member of the class and has not previously been characterised electrophysiologically. Because the transient outward current is itself a validated determinant of the BrS substrate (gain-of-function variants in the *KCND3*-encoded Kv4.3 channel that carries I_to_ cause BrS16), we asked how AR-788 behaves in a human BrS model. Using right-ventricular *SCN5A*-truncation cardioids, we report that AR-788’s dominant, reproducible action in this system is not on I_Na_ but on I_to_: it suppresses the pathologically enhanced transient outward current in a concentration-dependent manner while sparing the sodium current. This establishes AR-788 as a candidate Ito-directed agent for Brugada syndrome and provides a human proof-of-concept for correcting the repolarizing-current excess that defines the disease substrate.

## Results

### Study design and model provenance

We differentiated three-dimensional cardioids from a wild-type (WT) control hiPSC line (H166 WTC09) and from two Brugada syndrome lines carrying *SCN5A* variants: Br694, which bears a frameshift variant (p.A665Gfs*16) introducing a premature stop codon, and MSB01, which bears the missense variant p.N1722D. Differentiation followed an adapted version of the Schmidt protocol^13^, with a combination of factors directing a right-ventricular identity (Fig. 1a,b). Across twelve independent differentiation runs, the WT and Br694 lines grew and beat reproducibly (4/4 and 3/3 runs, respectively), whereas both independent MSB01 clones tested (C37 and C38) failed in every attempt (0/5): cardioids progressively fragmented and ceased beating during differentiation (Supplementary Fig. 1). The missense line therefore contributes no data to this study, and all subsequent comparisons are between the WT line and the *SCN5A* frameshift line. Cardioids were used intact for optical mapping at day 14.5–16.5, or dissociated at day 15.5 and re-plated in two dimensions for approximately one week before manual patch-clamp at day 21.5–23.5 (Fig. 1b).

**Figure 1.**
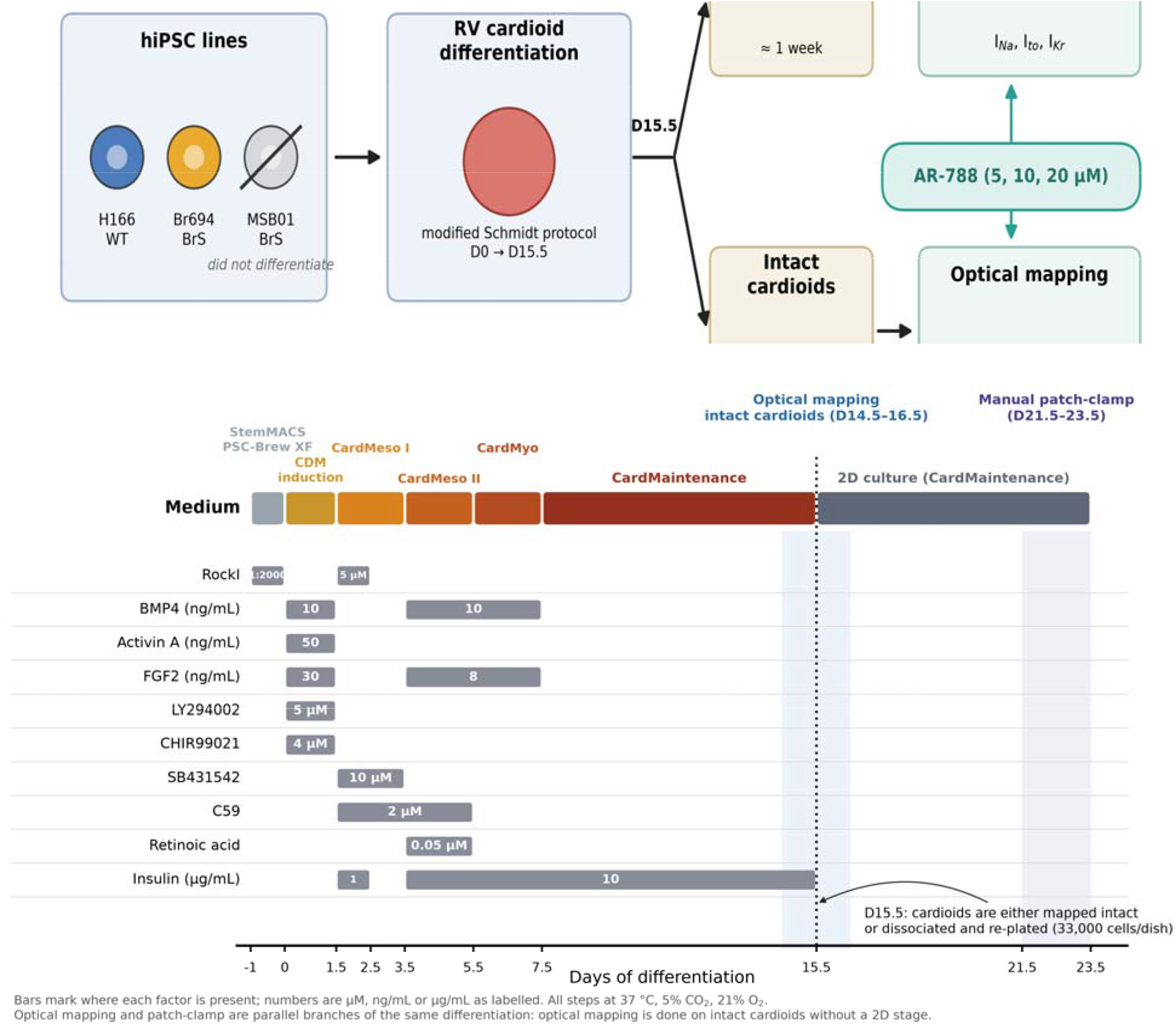
Study design and differentiation protocol. (a) Schematic overview of the hiPSC lines (WT, H166 WTC09; BrS Br694, SCN5A p.A665Gfs*16; BrS MSB01, SCN5A p.N1722D, which failed to differentiate), cardioid differentiation, dissociation to 2D culture, and the single-cell patch-clamp (I_Na_, I_to_, I_Kr_) and optical-mapping (action potential, calcium transient) read-outs, with AR-788 (AR) applied to both. (b) Day-resolved differentiation protocol, adapted from Schmidt et al._^13^_. Bars mark the interval over which each factor is present.Optical mapping and patch-clamp are parallel branches of the same differentiation, not sequential steps.

### BrS cardioids reproduce the sodium-channel loss-of-function phenotype

Cardioids were dissociated at day 15.5 and studied by manual patch-clamp, with sodium currents recorded on perfused cells at 35–37 °C. Peak I_Na_ density at −20 mV was reduced by ~60% in BrS relative to WT cardiomyocytes (−40.67 vs −100.6 pA pF−1; p = 0.026), and the current–voltage relationship was depressed across the activation range (Fig. 2a,b; Supplementary Table 1). The voltage dependence of activation was comparable between genotypes with a modest depolarizing tendency in BrS, and the slope factor (k) of the peak-current relationship was slightly increased (Fig. 2c; Supplementary Table 1). These data confirm a loss-of-function sodium-channel phenotype consistent with BrS^9,10^.

**Figure 2.**
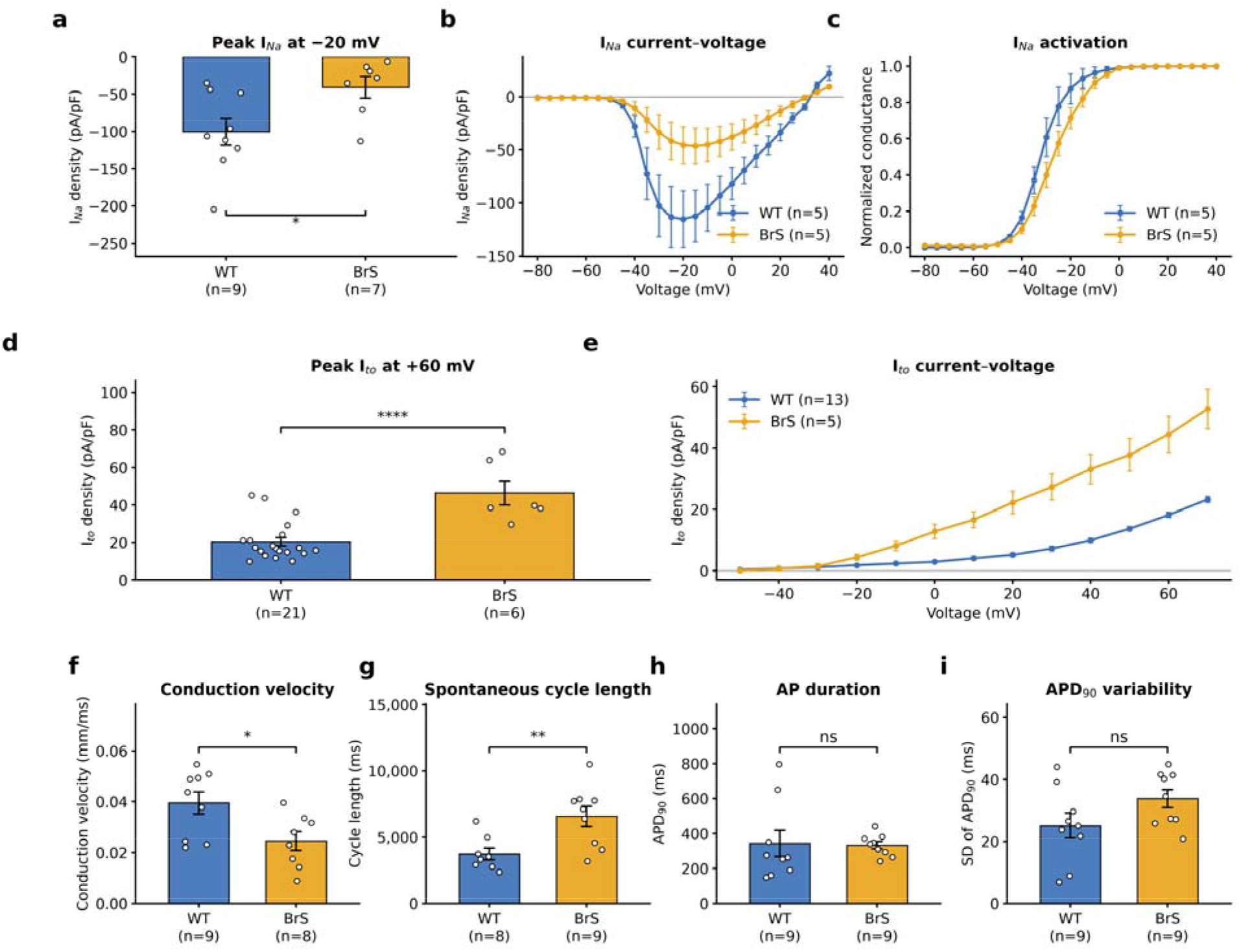
Electrophysiological phenotype of BrS cardioids. (a) Peak INa density at −20 mV in WT and BrS hiPSC-derived cardiomyocytes. (b) Mean INa current–voltage relationships. (c) Voltage dependence of INa activation. (d) Peak Ito density at +60 mV. (e) Mean Ito current–voltage relationships. (f) Conduction velocity, (g) spontaneous cycle length, (h) action-potential duration at 90% repolarisation (APD90) and (i) beat-to-beat SD of APD90 measured by optical mapping of WT and BrS cardioids. Bars and points are mean ± SEM. Circles (⍰) are individual cells (a, WT n = 9, BrS n = 7; d, WT n = 21, BrS n = 6) or cardioids (f–i; n as indicated on the axes); curves are mean ± SEM of n = 5 cells per genotype (b, c) or WT n = 13 and BrS n = 5 cells (e). *p < 0.05, **p < 0.01, ****p < 0.0001, ns = not significant.

### The transient outward current is markedly enhanced in BrS cardioids

In contrast to the reduced sodium current, the transient outward potassium current was substantially larger in BrS cardioids. Peak I_to_ density at +60 mV was more than doubled relative to WT (46.38 vs 20.40 pA pF−1; +127%; p < 0.0001), with enhancement across the entire suprathreshold voltage range (Fig. 2d,e; Supplementary Table 1). An I_Na_ deficit combined with an I_to_ excess is the imbalance predicted to accentuate the epicardial phase-1 notch and to favour loss of the action-potential dome^6,7,16^.

### Tissue-level phenotype: slowed conduction and abnormal spontaneous activity

Optical mapping of intact cardioids revealed a tissue-level correlate of the ionic phenotype^17^. Conduction velocity was reduced in BrS relative to WT (0.02458 vs 0.03949 mm/ms; −38%; p = 0.0216), consistent with reduced I_Na_, and the spontaneous cycle length was markedly prolonged (6571 vs 3734 ms; +76%; p = 0.0073). Action-potential amplitude and mean action-potential duration at 90% repolarisation (APD90) did not differ significantly, although beat-to-beat APD90 variability trended higher in BrS (Fig. 2f–i; Supplementary Table 1).

### The disease shifts the phase-1 current balance 5.6-fold

Relative to the WT reference, BrS cardioids display a coordinated multi-parameter shift, reduced I_Na_, enhanced I_to_, slowed conduction, and prolonged, more variable spontaneous activity, that together define the disease substrate in this model (Fig. 3a). The repolarization hypothesis predicts that the ratio of the two currents, not either current alone, sets the balance. Expressed as the cohort-level ratio of repolarizing to depolarizing current, I_to_/|I_Na_| rose from 0.20 in WT to 1.14 in BrS cardiomyocytes, a 5.6-fold shift in the balance that governs the phase-1 notch (Fig. 3b). Because I_to_ and I_Na_ were recorded in separate cells, this index is a group-level descriptor rather than a paired measurement.

**Figure 3.**
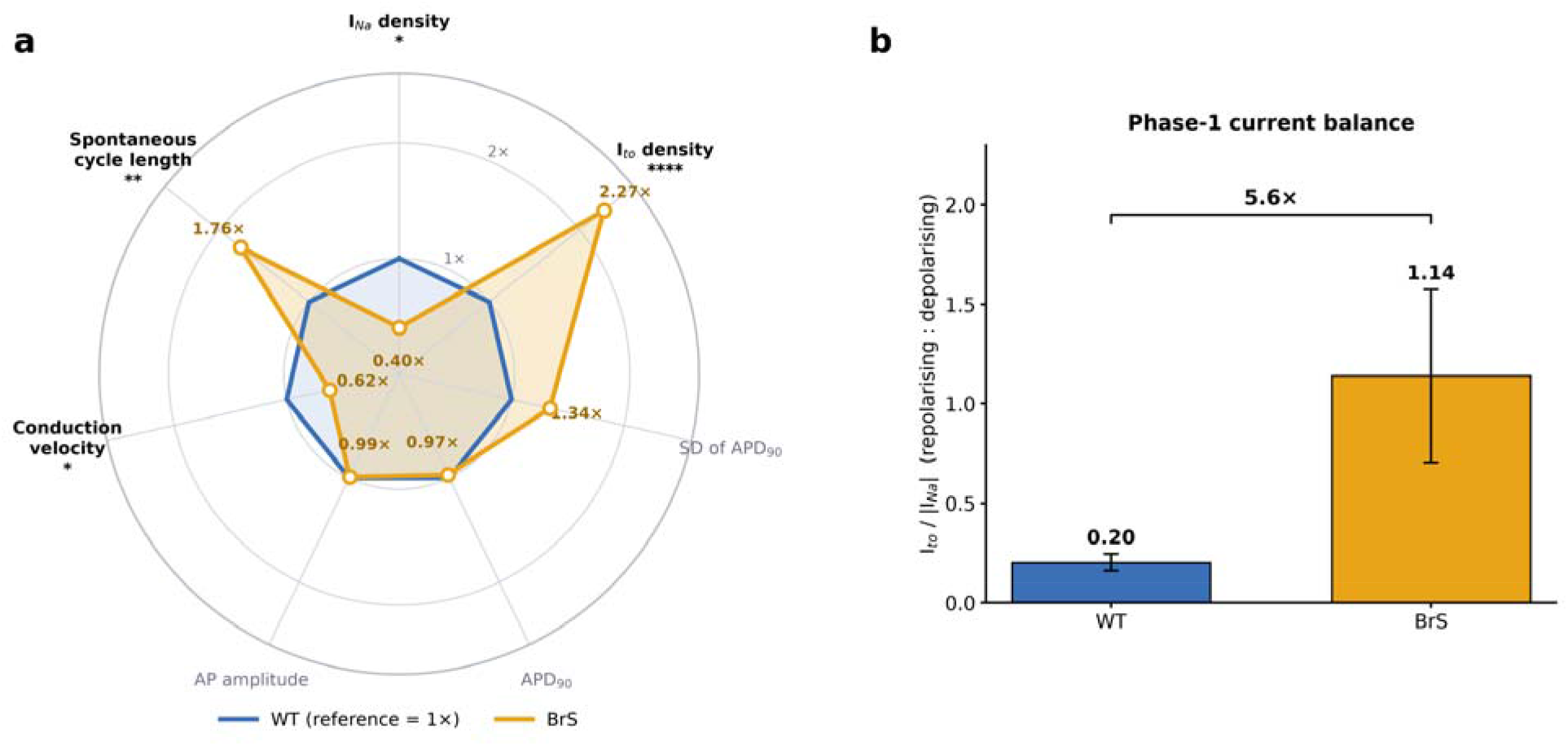
Integrated Brugada phenotype. (a) Radar summary of every measured baseline parameter in BrS cardioids expressed as a ratio to the WT reference (set to 1×). Bold axis labels reach p < 0.05. (b) Cohort-level ratio of the two currents that set the phase-1 balance, Ito/|INa|, in WT and BrS cardiomyocytes; error bars are propagated SEM. Ito and INa were recorded in different cells, so the ratio is a group-level index, not a paired measurement.

### AR-788 suppresses I_to_ and normalises it in BrS cardiomyocytes

Exposure to AR-788 (AR) at 10 µM reduced peak I_to_ density at +60 mV by 49% in BrS cardiomyocytes (42.53 → 21.71 pA pF−1; p = 0.0218) and by 35% in WT cardiomyocytes (21.48 → 13.80 pA pF−1; p = 0.0005), with suppression across the depolarized I–V range in both genotypes (Fig. 4a–d; Supplementary Table 2). Because AR lowered I_to_ in both BrS and WT cells, its action is on the current itself rather than on a BrS-specific channel population; the larger absolute effect in BrS reflects the larger starting I_to_. Basal values here are those of the cells that completed the AR protocol (BrS n = 4, WT n = 9) and therefore differ from the phenotyping cohort (Fig. 2d).

**Figure 4.**
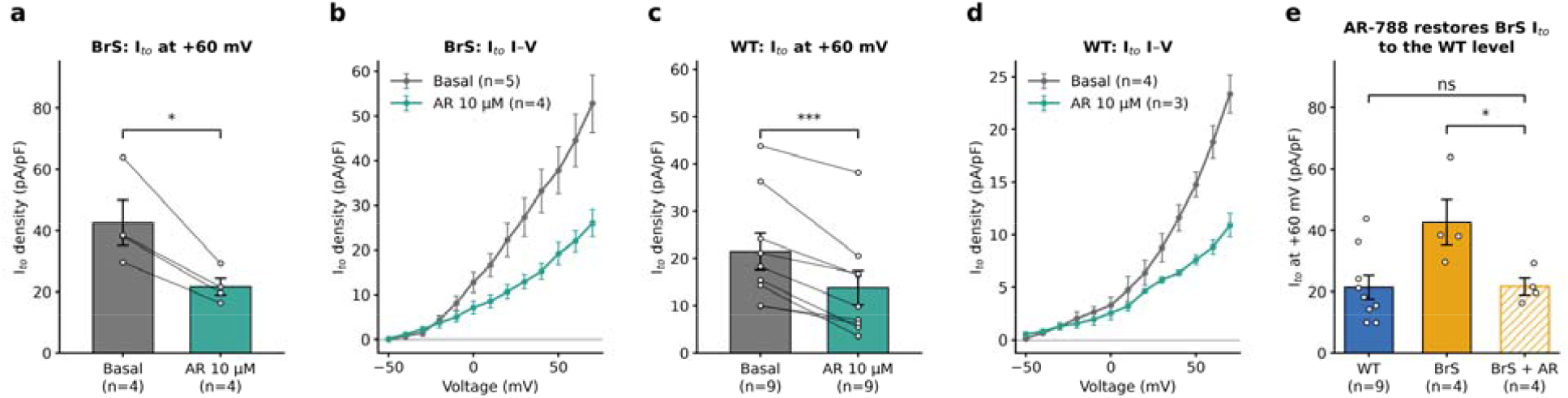
AR-788 inhibits Ito. (a) BrS Ito density at +60 mV before/after AR 10 µM and (b) corresponding I–V relationships. (c) WT Ito density at +60 mV before/after AR 10 µM and (d) I–V relationships. Bars and points are mean ± SEM; lines connect the paired recordings of each cell (n as indicated in each panel); *p < 0.05, ***p < 0.001. (e) Peak Ito at +60 mV in WT, untreated BrS and AR-treated BrS cardiomyocytes; * refers to BrS basal versus BrS + AR (paired) and ns to BrS + AR versus WT (p = 0.9716).

Treated BrS cardiomyocytes were statistically indistinguishable from untreated WT cardiomyocytes (21.71 vs 21.48 pA pF−1; p = 0.9716; n = 4 BrS, 9 WT; Fig. 4e), so at 10 µM AR-788 returned the current to the wild-type range rather than only attenuating it. The consequence for the phase-1 current balance is described below.

### I_to_ inhibition is concentration-dependent

In WT cardiomyocytes, AR-788 inhibited I_to_ concentration-dependently. Normalized peak I_to_ fell to 0.88, 0.70 and 0.58 of baseline at 5, 10 and 20 µM (12.3%, 29.7% and 42.4% inhibition; 10 and 20 µM: p = 0.0004 and p < 0.0001; Fig. 5; Supplementary Table 3). A Hill fit constrained to complete block returned an extrapolated IC50 of 25.4 ± 3.2 µM with a Hill coefficient of 1.09 ± 0.17, consistent with a single-site interaction of moderate potency; fixing the Hill coefficient to unity gave a closely similar IC50 of 26.9 ± 1.9 µM. A three-parameter fit with maximal block left free is degenerate on three informative points and was not used. Because the IC50 lies beyond the highest concentration tested, it remains an extrapolation and should be confirmed over a wider concentration range.

**Figure 5.**
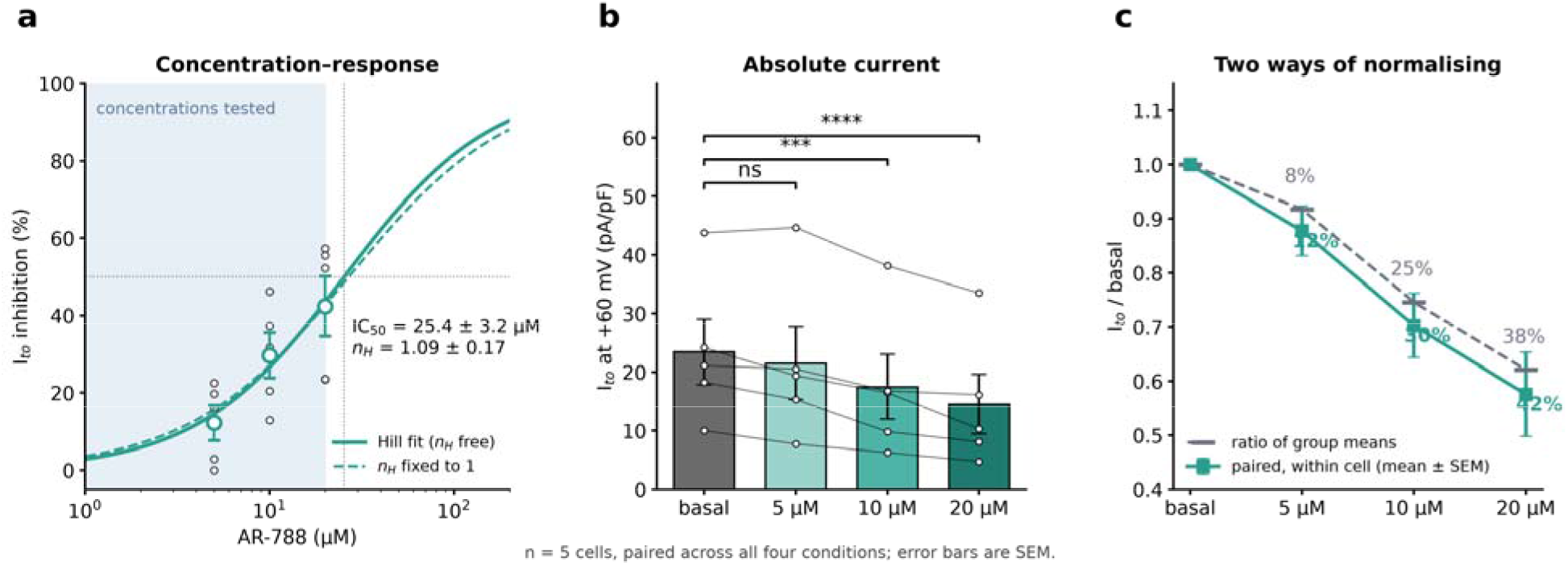
Concentration dependence of Ito inhibition (WT cardiomyocytes). (a) Percent Ito inhibition versus AR concentration with a Hill fit constrained to complete block (solid, Hill coefficient free; dashed, Hill coefficient fixed to 1). Shading marks the range of concentrations actually tested. (b) Absolute Ito density at +60 mV across concentrations; n = 5 cells paired across all four conditions. (c) The two normalisations used in this study, which differ. Error bars are SEM and circles (⍰) are individual cells. ns, not significant; ***p < 0.001, ****p < 0.0001. The full current–voltage series at 20 µM was available for one cell only; all concentration–response values shown here are peak Ito at +60 mV in five cells.

Two normalisations appear in this dataset and they are not interchangeable. The percent inhibition quoted above is the mean of each cell’s own basal-normalised ratio across the paired series (n = 5 cells carried through all four conditions); the ratio of the group-mean current densities gives systematically smaller values (8.3%, 25.4% and 37.9%). Both are reported in Supplementary Table 3, and the paired values are the ones quoted in the text and used for the Hill fit (Fig. 5c).

### AR-788 spares the sodium current

AR-788 did not reduce I_Na_. At 10 µM, peak I_Na_ density at −20 mV was unchanged in BrS cardiomyocytes (−15.74 vs −16.39 pA pF−1; p = 0.3703) and in WT cardiomyocytes (−37.49 vs −33.34 pA pF−1; p = 0.2101), and the full I–V relationships were superimposable before and after exposure in both genotypes (Fig. 6a–d; Supplementary Table 2). AR-788 did not rescue the sodium current, so the original therapeutic hypothesis for the ARumenamide series (stabilisation of Nav1.5 availability in a loss-of-function setting) was not met in this model. It also did not depress I_Na_ further, which matters because additional sodium-channel block would aggravate Brugada syndrome.

**Figure 6.**
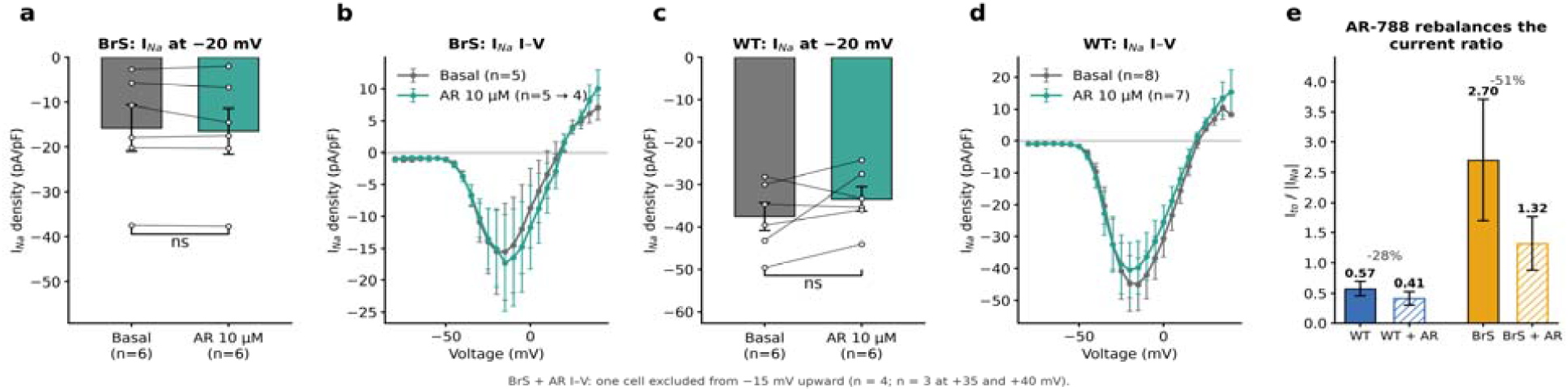
AR-788 does not reduce I_Na_. (a) BrS I_Na_ density at −20 mV and (b) I–V relationships before/after AR 10 µM. (c) WT INa density at −20 mV and (d) I–V relationships. Bars and points are mean ± SEM; lines connect the paired recordings of each cell. Density: n = 6 cells per genotype and condition. I–V curves: BrS n = 5 (basal) and n = 5 (AR 10 µM; n = 4 from −15 mV upward and n = 3 at +35 and +40 mV, one cell being excluded from the mean), WT n = 8 (basal) and n = 7 (AR 10 µM). (e) Ratio of the group-mean Ito to |INa| (external Na+ 10 mM), with propagated SEM; Ito and INa were recorded in different cells, so the ratio is a group-level index. ns = not significant.

Note that absolute I_Na_ densities in the AR-788 experiments are smaller than those in the phenotyping experiments because the external Na+ concentration was lowered from 20 to 10 mM for the second part of the study (see Methods). Within-experiment comparisons are unaffected, but absolute densities should not be compared between Supplementary Table 1 and Supplementary Table 2.

### AR-788 rebalances the phase-1 current ratio

Because the phase-1 notch is set by the balance between Ito and INa rather than by either current alone, we also expressed the effect of AR-788 as the ratio of group-mean Ito to |INa| (Fig. 6e). The AR-788 experiments used 10 mM external Na+, so absolute values are not comparable with Fig. 3b. In BrS cardiomyocytes AR-788 lowered the index by 51% (2.70 → 1.32), against a 28% fall in WT cells (0.57 → 0.41), so that the treated BrS ratio moved towards, but did not reach, the WT baseline. Because Ito and INa were recorded in different cells, the index is a group-level descriptor and its error bars are propagated SEM.

### Cardiac safety: effect on IKr

Because I_to_-directed agents must not unduly impair rapid delayed-rectifier repolarization^18^, we assessed IKr (carried by hERG, also known as Kv11.1)^19^, normalizing to the maximal block produced by BeKm-1 (ref. 20). AR-788 10 µM reduced the peak IKr tail current by 40% (2.85 → 1.70 pA pF−1; p = 0.0035; n = 7), against a residual current in BeKm-1 of 1.02 pA pF−1. The voltage dependence of activation was unchanged (V1/2 −33.75 vs −32.65 mV; p = 0.58) as was its slope (p = 0.33), and the I–V relationship was depressed without a change in shape (Fig. 7a–c; Supplementary Table 2). A 40% reduction in I_Kr_ amplitude at a concentration above the anticipated therapeutic range is a safety liability. Formal proarrhythmia profiling, including CiPA-style assessment and a full hERG concentration–response, will be required before this chemotype advances. The absence of any gating shift suggests open-channel block rather than trafficking or gating modification, which narrows the experiments needed to characterise it.

**Figure 7.**
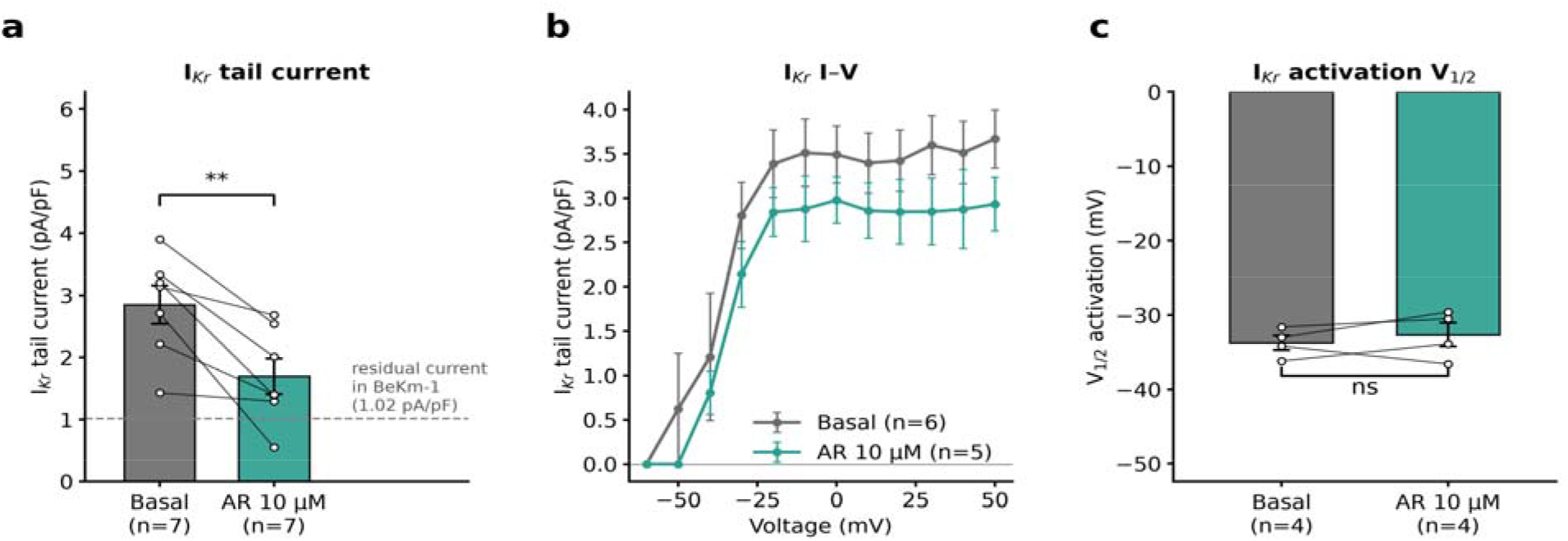
Effect of AR-788 on IKr. **(a) Peak IKr tail current** before/after AR 10 µM, normalized to the maximal BeKm-1– sensitive block; the dashed line marks the residual current in BeKm-1. (b) IKr I–V relationships. (c) Activation V1/2. Bars and points are mean ± SEM; lines connect paired recordings of each cell (a, n = 7; b, n = 6 basal and n = 5 AR; c, n = 4). **p < 0.01, ns = not significant.

### Tissue-level effects on action potentials and calcium transients

At the tissue level, optical mapping showed no significant change in any measured parameter after AR 10 µM, in either genotype. Consistent with the absence of an effect on I_Na_, conduction velocity was unchanged in WT (0.0313 → 0.0262 mm ms−1; p = 0.2022; n = 4) and in BrS cardioids (0.0164 → 0.0185 mm ms−1; p = 0.6449; n = 6). APD90 was likewise unchanged (WT 291 → 335 ms, p = 0.0996; BrS 189.3 → 195 ms, p = 0.9268). Spontaneous cycle length shortened in both genotypes without reaching significance (WT 8930 → 4679 ms, p = 0.1041; BrS 4293 → 3007 ms, p = 0.1941). Beat-to-beat APD90 variability fell in BrS cardioids (38.51 → 28.92 ms; p = 0.1142) while rising in WT (25.93 → 32.72 ms; p = 0.4203), a genotype-dependent direction that is consistent with a reduction of repolarization heterogeneity where I_to_ is pathologically elevated, but which does not reach significance and must be treated as hypothesis-generating. Calcium-transient recordings in BrS cardioids gave the same picture: conduction velocity 0.00880 → 0.00562 mm/ms (p = 0.0955), cycle length 3385 → 3232 ms (p = 0.8389), amplitude 5.989 → 5.873 (p = 0.7066), calcium-transient duration at 80% repolarisation (CaD80) 640 → 629.5 ms (p = 0.8601) and SD of CaD80 72.84 → 63.50 ms (p = 0.1469) (Fig. 8a–d; Supplementary Table 4).

**Figure 8.**
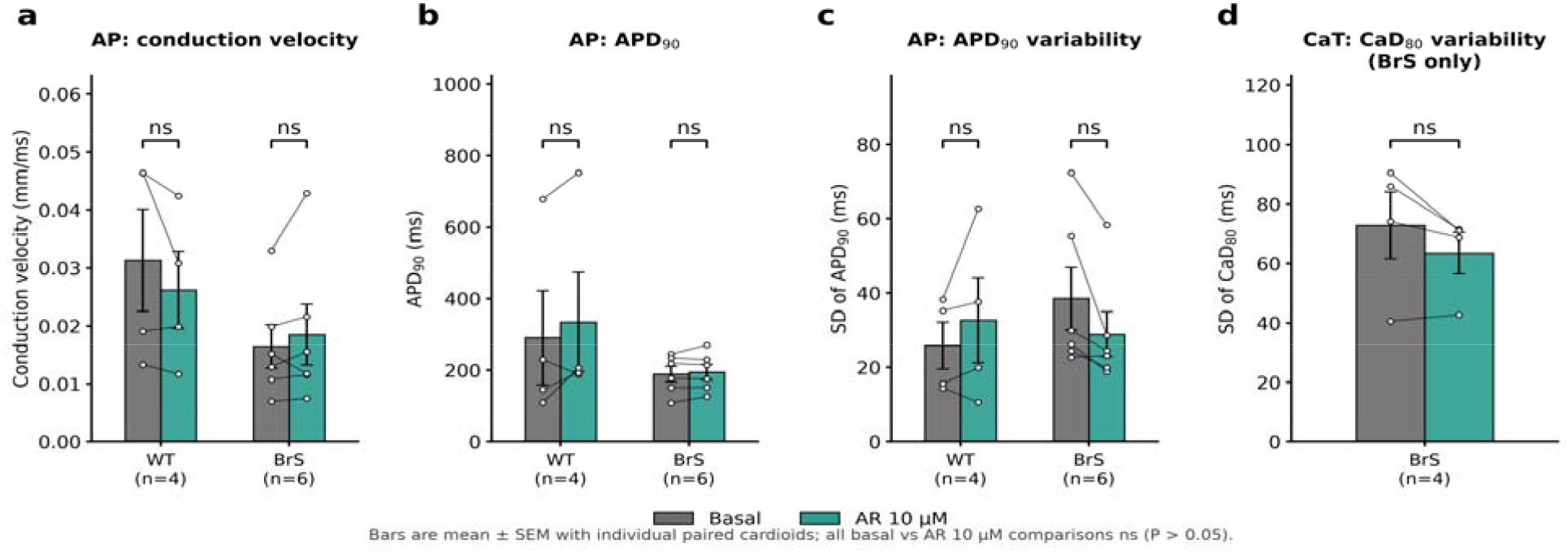
Tissue-level effects of AR-788 in WT and BrS cardioids. (a) Conduction velocity, (b) APD90, (c) SD of APD90 (action-potential mapping) and (d) SD of CaD80 (calcium-transient mapping) before/after AR 10 µM. Bars are mean ± SEM; circles (⍰) joined by lines are individual cardioids (WT n = 4, BrS n = 6, calcium transients n = 4). ns = not significant; small n (see Methods).

These multicellular recordings were technically constrained: of 17 cardioids surveyed, only 4 beat sufficiently for calcium-transient analysis, and action-potential maps were analysable in four WT and six BrS cardioids. Among the preparations that could not be analysed, some failed to beat at all while others beat with early afterdepolarizations (EADs) already present before any drug exposure; EADs preclude the measurement of conduction and of calcium-transient kinetics, which is why those recordings were excluded. We report this spontaneous arrhythmic activity because it is a phenotypic observation in its own right, while noting that it was not quantified systematically and that the denominator above was not designed as an arrhythmia endpoint. The tissue-level data are therefore underpowered, preliminary and hypothesis-generating; the single-cell current measurements constitute the primary evidence of this study.

### An integrated view of disease and drug

Taken together, the measured parameters fall into three groups (Fig. 9c). The disease signature is dominated by a 60% loss of I_Na_ and a 127% gain of I_to_, with slowed conduction and prolonged cycle length following. The single-cell pharmacology of AR-788 is confined to the potassium currents: I_to_ falls by 35–49% and the IKr tail current by 40%, while I_Na_ moves by 11% or less, within the range expected from recording variability. The multicellular effects are uniformly small and non-significant. The compound’s profile is therefore the reverse of its design hypothesis (Fig. 9a,b).

**Figure 9.**
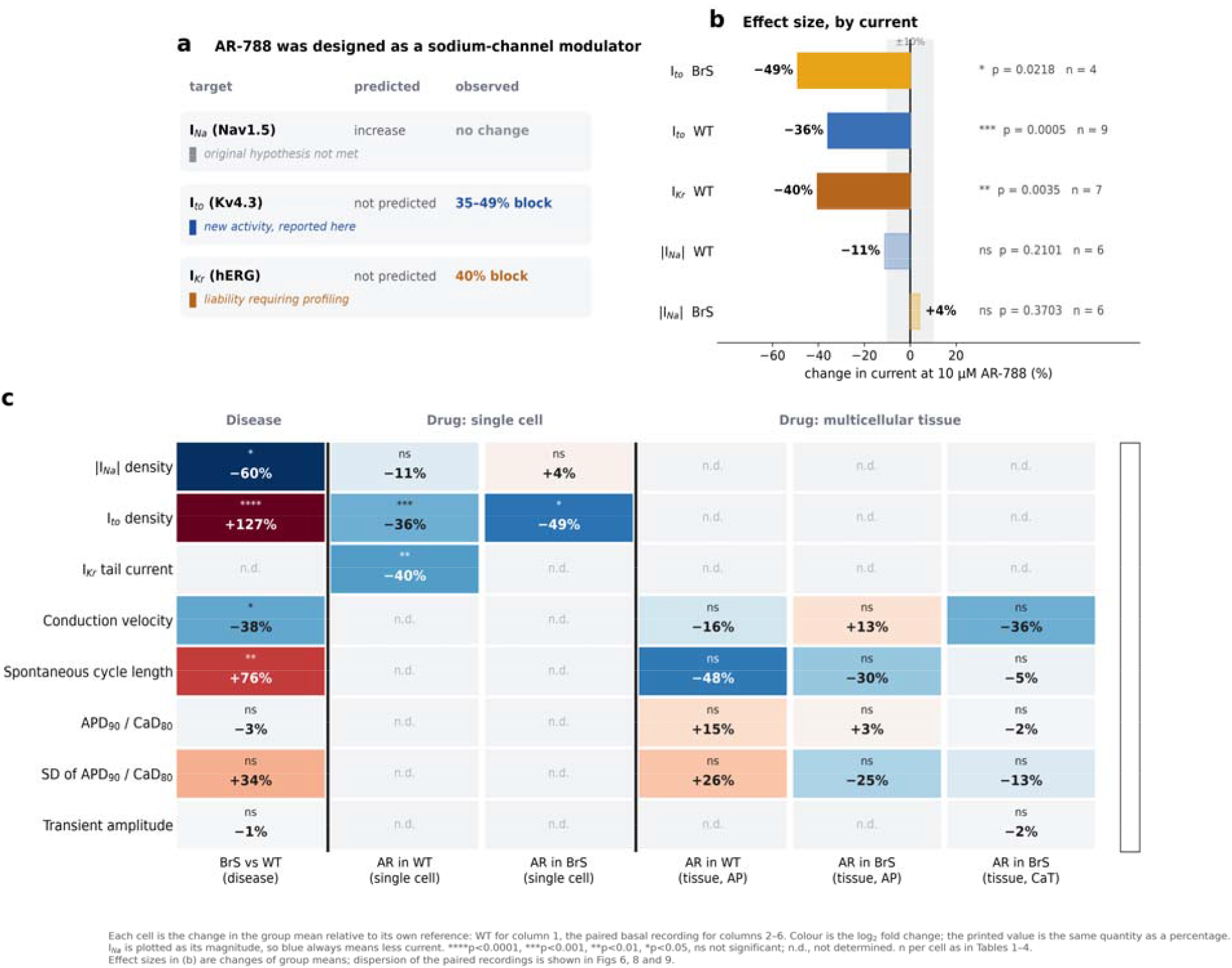
Pharmacological reframing and integrated summary. (a) Predicted versus observed activity at each of the three currents assayed. (b) Effect size at 10 µM AR-788, by current; solid bars reach p < 0.05, faded bars do not, and shading marks a ±10% band around zero. (c) Every comparison in the study on one scale: colour is the log2 fold-change relative to wild-type (first column) or to the paired basal recording (remaining columns); the printed value is the same quantity as a percentage. INa is plotted as its magnitude, so blue always means less current. n.d., not determined. Effect sizes in (b) are changes of group means; the dispersion of the paired recordings is shown in Figs 4, 6 and 7.

### Proposed mechanism

Together, these data support a model in which AR-788 re-balances the BrS current profile primarily by lowering the pathologically enhanced I_to_ while sparing I_Na_, partially restoring the epicardial action-potential dome and reducing the substrate for phase-2 reentry (Fig. 10a–c). The chemical identity of AR-788 and its provisional site of action are summarized in Fig. 10d,e.

**Figure 10.**
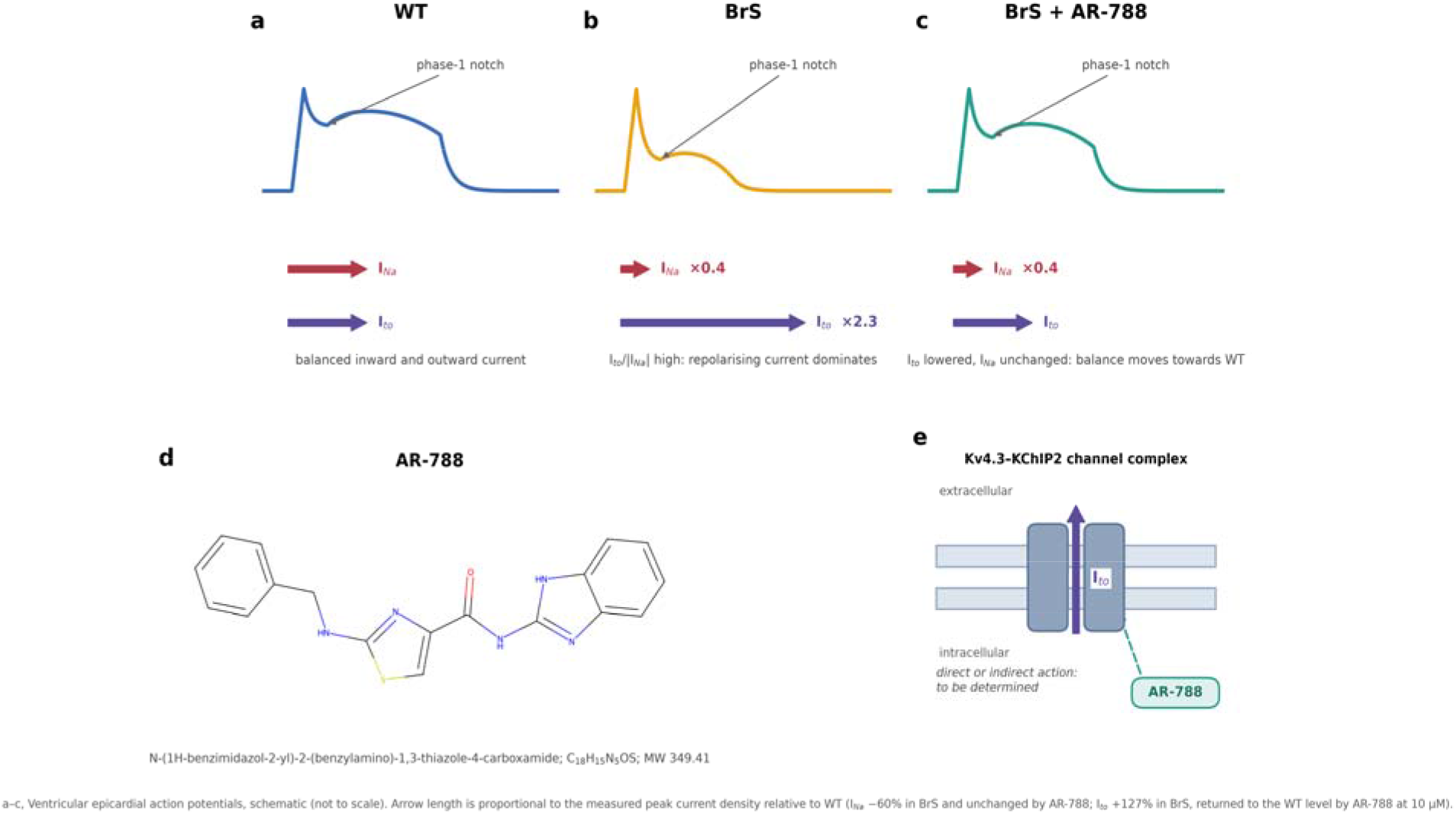
Proposed mechanism, structure of AR-788 and putative target. (a–c) Conceptual model of the ventricular epicardial action-potential notch in WT, BrS and BrS + AR-788. In BrS, reduced INa together with enhanced Ito exaggerates the phase-1 notch and favours loss of the dome; AR-788 lowers Ito, re-balancing depolarizing and repolarizing currents and partially restoring the dome. Arrow length is proportional to the measured peak current density relative to WT: INa is 60% lower in BrS and unchanged by AR-788, whereas Ito is 2.3-fold (+127%) higher in BrS and returns to the WT level with AR-788 at 10 µM. (d) Chemical structure of AR-788 (N-(1H-benzimidazol-2-yl)-2-(benzylamino)-1,3-thiazole-4-carboxamide; C18H15N5OS; MW 349.41). (e) Kv4.3–KChIP2 channel complex carrying Ito; whether AR-788 acts on this channel directly or indirectly remains to be determined.

## Discussion

Using human iPSC-derived cardioids patterned to a right-ventricular identity and carrying an *SCN5A* frameshift variant, we show that the Brugada phenotype in this model comprises not only a reduced sodium current but a pronounced excess of the transient outward potassium current, and that AR-788, a member of a class framed primarily around Nav1.5 modulation, acts here principally as a concentration-dependent inhibitor of Ito while sparing INa. This extends the Ito-directed pharmacology of the ARumenamide class and offers a human proof-of-concept for correcting the repolarizing-current excess central to the BrS substrate.

The therapeutic logic follows directly from the mechanistic model of BrS^5,6^. Because the arrhythmogenic substrate arises from an imbalance between reduced inward and preserved or enhanced outward currents in the early action potential, an agent that lowers Ito can restore balance even without repairing the sodium-channel lesion, which is the accepted basis for the efficacy of quinidine^4,7^. Our data position AR-788 as a chemically distinct I_to_-directed candidate, and it did not further reduce I_Na_, which matters because additional sodium-current block would aggravate the phenotype. The treated BrS transient outward current was statistically indistinguishable from the untreated WT current (p = 0.9716). The compound did not simply blunt Ito; it restored it to the wild-type range at a concentration that left the sodium current intact.

The right-ventricular identity of the model matters for interpretation. The Brugada substrate is regional: it resides in the right-ventricular outflow tract, where the epicardial transient outward current is largest and the phase-1 notch consequently deepest^6,7^. The directed differentiation protocol13 produced cardioids of right-ventricular character, so the I_to_ excess reported here was measured in the chamber in which the disease expresses itself, rather than in an unspecified ventricular-like population. Chamber identity is therefore, we argue, a requirement for any I_to_-directed pharmacology tested in a BrS model.

The missense line failed to differentiate, and we report this in full. Both independent MSB01 clones (p.N1722D) fragmented and ceased beating across five differentiation runs, while the WT and frameshift lines succeeded in all seven of theirs. Whether this reflects a genuine biological consequence of the variant, a clone-specific artefact, or a differentiation-protocol incompatibility cannot be resolved from these data, and we make no claim either way; it does mean the study rests on a single BrS genotype.

Several limitations temper interpretation. First, the single-cell findings, while internally consistent and statistically significant for I_to_, rest on one successfully differentiating BrS line; a second, independent BrS line, ideally isogenic-corrected, is needed to establish generality^9^. Second, the tissue-level optical-mapping and calcium-transient experiments were underpowered, so the action-potential-level consequences of Ito suppression remain to be demonstrated convincingly; the genotype-dependent direction of the APD90-variability change is suggestive but did not reach significance. Third, the concentration–response places the IC50 beyond the highest tested concentration (25.4 ± 3.2 µM extrapolated); potency, selectivity and the molecular target of I_to_ inhibition (the Kv4.3–KChIP2 channel complex versus an indirect action) require dedicated experiments in a heterologous system^16^. Fourth, the 40% reduction of the IKr tail current at 10 µM, although unaccompanied by any gating shift, is a substantive liability and mandates formal proarrhythmia assessment; we note that both the Ito and the hERG effects were measured at the same concentration, so the therapeutic window between the intended and the off-target potassium-channel actions is presently unknown and is the first thing a follow-up study should establish. Fifth, recordings were made in an open-label, non-randomized design, as is standard for manual patch-clamp. Finally, hiPSC-cardiomyocyte immaturity is a general constraint of the platform^21^.

Despite these limitations, a defined small molecule reproducibly and dose-dependently suppressed the enhanced I_to_ of human right-ventricular BrS cardiomyocytes and returned it to the wild-type range without reducing I_Na_. This provides a chemically tractable starting point for Brugada pharmacotherapy and motivates target deconvolution, isogenic validation, hERG counter-screening and in vivo testing.

## Methods

### hiPSC lines

Three human induced pluripotent stem cell lines were used. The WT line, H166 WTC09 (ITXi002-A; Cellosaurus CVCL_B5QD), was generated from primary dermal fibroblasts, has been characterised previously and is routinely used by our team22–24. The two Brugada syndrome lines were generated from blood samples of patients with Brugada syndrome: Br694, carrying a frameshift variant in *SCN5A* (p.A665Gfs*16) that introduces a premature stop codon, and MSB01, carrying the missense variant *SCN5A* p.N1722D. Two independent clones of MSB01 (C37 and C38) were tested.

### Cardioid differentiation

Cardioids were generated using an adapted version of the protocol published by Schmidt et al.13. All culture steps were performed at 37 °C in 5% CO2 and 21% O2. On day −1, hiPSCs were seeded into a 24-well plate pre-coated with Matrigel hESC-Qualified Matrix (0.05 mg mL−1) at 40,000 cells per well in StemMacs PSC-Brew XF human Medium (Miltenyi Biotec #130-127-865) supplemented with ROCK inhibitor (RockI, 1:2000, Tocris #1254). On day 0, mesodermal differentiation was induced by replacing the medium with Cardiac Differentiation Medium (CDM; 50% IMDM (Gibco #21980065), 50% DMEM F12 Nut-Mix (Gibco #31765068), 4 mg mL−1 BSA (Sigma-Aldrich #A9418; 0.22 µm filtered), 1% lipid mix (Gibco #11905031), 0.004% monothioglycerol (Sigma-Aldrich #M6145) and 15 µg mL−1 transferrin (Roche #10652202001)) supplemented with BMP4 (10 ng mL−1), Activin A (50 ng mL−1), FGF2 (30 ng mL−1), LY294002 (5 µM) and CHIR99021 (4 µM); this induction medium was maintained for 36–40 h. On day 1.5, cells were dissociated with 1× TrypLE (Gibco #12604-013) and reseeded at 20,000 cells per well in a 96-well ultra-low-attachment plate (Corning #7007) in cardiac mesodermal patterning medium I (CardMeso I; CDM containing SB431542 (10 µM) and C59 (2 µM)) supplemented with RockI (5 µM) and with insulin (1 µg mL−1); the plate was centrifuged for 4 min at 200g to induce aggregation. On day 2.5 the medium was replaced with CardMeso I without RockI or insulin. On days 3.5 and 4.5 the medium was replaced with cardiac mesoderm patterning medium II (CardMeso II) supplemented with insulin (10 µg mL−1), retinoic acid (0.05 µM), FGF2 (8 ng mL−1), BMP4 (10 ng mL−1) and C59 (2 µM). Cardiomyocyte specification was initiated on day 5.5 by replacing the medium with cardiomyocyte specification medium (CardMyo) at the same insulin, FGF2 and BMP4 concentrations as on days 3.5–4.5, renewed on day 6.5. From day 7.5, cardiomyocytes were maintained in CardMaintenance medium (CDM supplemented with insulin, 10 µg mL−1), replaced every 48 h until day 15.5. Catalogue details of the molecules used are given in Supplementary Table 5.

### Differentiation efficiency

Differentiation efficiency was assessed for each line across twelve independent runs. The WT line (H166 WTC09, 4/4 runs) and the BrS *SCN5A* frameshift line (Br694 clone C02, 3/3 runs) both showed good cardioid growth with an adequate proportion of beating cardioids. The BrS *SCN5A* missense line (MSB01) did not differentiate successfully in any of five runs across two independent clones (C37 and C38): cardioids progressively fell apart and ceased beating during differentiation. MSB01 was therefore excluded from all analyses (Supplementary Fig. 1).

### Cardioid dissociation for patch-clamp

D15.5 cardioids were pooled into a tube and washed 3 times with PBS, then transferred into a MW6 well with 1mL of warm TrypLE 10X (Gibco #1217701) and incubated for 5min at 37°C. 400 to 700µL (according to the number of cardioids) of warm CDM with 10mg/mL collagenase (Gibco #17101015) prepared extemporaneously were added to the well and the cardioids were incubated for 5min at 37°C. With gentle flushing and extra incubation if needed, cardioids were dissociated into single cells. The reaction was stopped with 10 to 15mL of warm CDM-BSA 1% (Sigma #A9418) while sieving the cell solution (70µm, Falcon #352350). After centrifugation for 5min at 260g at RT, the cell pellet was resuspended in 0.5-1mL CDM and used to seed the cells for subsequent patch-clamp experiments.

35mm petri dishes (NUNC #055055) were coated with 0.05mg/mL Matrigel hESC-Qualified Matrix in DMEM F12 and seeded with 33k cells in CardMaintenance. The medium was changed with CardMaintenance the following day then every other day. Patch-clamp measurements were conducted from days 6 to 8 after dissociation i.e. D21.5-23.5 of the differentiation.

### Manual patch-clamp electrophysiology

Ionic currents were recorded by manual whole-cell patch-clamp on cells between day 21.5 and day 23.5, following dissociation of day-15.5 cardioids and re-plating in two dimensions. The experiment was not randomized and was conducted open-label with respect to the condition of the cells under study. Sodium current recordings were performed on perfused cells with the temperature maintained between 35 and 37 °C. Recordings used a VE-2 amplifier (Alembic Instruments, Montreal, QC, Canada), an Axon CNS Digidata 1440A analog-to-digital converter and Clampex 10.3 software (Molecular Devices, Sunnyvale, CA, USA). Micropipettes of 1.2–2.4 MΩ resistance were fabricated from borosilicate capillaries on a Sutter Instruments P-97 puller. Signals were low-pass filtered at 10 kHz. Cells were immersed in a Tyrode’s solution containing (in mM): NaCl 130, KCl 4, NaH2PO4 1.2, MgSO4·7H2O 1.2, HEPES 10, glucose 5 and CaCl2 0.001; pH adjusted to 7.2 with NaOH. A current-specific external control solution was applied locally during recordings. AR-788 was added to an external solution otherwise identical to the control solution and applied to the same cell for a second series of recordings. Series resistance was compensated; after leak subtraction, all current measurements were normalized to membrane capacitance. Activation curves were fitted with a Boltzmann function, Y = Ymax/(1 + exp(−(x − V1/2)/k)), where Ymax is the maximum measured conductance, V1/2 the potential of half-maximal activation and k the slope factor. Data were analysed in Clampfit (Axon Instruments, Union City, CA, USA) and Prism 5 (GraphPad Software).

### Sodium current (INa)

Pipettes contained (in mM): NaCl 5, CsCl 45, HEPES 10, EGTA 1, Cs-gluconate 0.09 and MgATP 0.005; pH adjusted to 7.2 with CsOH. Two external solutions were used successively during the study. For the first part (characterization of the BrS phenotype versus WT): NaCl 20, CsCl 120, HEPES 10, glucose 10, mannitol 30, MgCl2 2 and CoCl2 2; pH adjusted to 7.4 with CsOH, with BeKm-1 (30 nM) subsequently added to inhibit hERG. For the second part (study of the effect of AR-788) a lower NaCl concentration was used: NaCl 10, with all other constituents unchanged. Because the transmembrane sodium gradient differs between the two solutions, absolute INa densities from the phenotyping experiments (Supplementary Table 1) and the AR-788 experiments (Supplementary Table 2) are not directly comparable; all comparisons reported are made within a single solution condition. INa at −20 mV was determined by applying depolarizing steps to −20 mV for 100 ms from a holding potential of −100 mV, with a 2 s interval between pulses. Current–voltage and activation relationships were obtained from a holding potential of −100 mV with steps from −80 to +40 mV in 5 mV increments for 50 ms, at 2 s intervals.

### Transient outward potassium current (Ito)

Pipettes contained (in mM): KCl 45, NaCl 5, HEPES 10, EGTA 1, K-gluconate 0.09 and MgATP 0.005; pH adjusted to 7.4 with KOH. The external solution contained (in mM): NaCl 136, KCl 4, HEPES 10, glucose 10, MgCl2 2 and CoCl2 2 with 0.5% BSA; pH adjusted to 7.4 with NaOH, with BeKm-1 (30 nM) and TTX (50 µM) subsequently added. Ito density was determined by applying depolarizing steps to +60 mV for 500 ms at 15 s intervals; Ito was isolated using a holding potential of −75 mV and a 30 ms prepulse to −50 mV to inactivate the sodium current. Current–voltage relationships were obtained from a holding potential of −75 mV with a 30 ms prepulse to −50 mV followed by steps from +70 to −50 mV in 10 mV decrements for 500 ms, at 15 s intervals.

### Rapid delayed-rectifier potassium current (IKr, hERG)

Pipettes contained (in mM): KCl 97, HEPES 10, EGTA 5, K-gluconate 0.045, K2ATP 1.5 and MgCl2 1; pH adjusted to 7.2 with KOH. The external solution contained (in mM): NaCl 130, KCl 4, NaH2PO4 1.2, MgSO4·7H2O 1.2, HEPES 10, glucose 5 and CaCl2 0.002 with 0.5% BSA; nifedipine (5 nM) and TTX (50 µM) were added at a later stage. IKr at −50 mV was determined from a holding potential of −50 mV followed by a prepulse to +10 mV and a step to −50 mV for 8 s, at 12.5 s intervals. The I–V relationship was obtained with a two-step protocol: from a holding potential of −50 mV, steps from −60 to +60 mV in 10 mV increments lasting 4 s were applied, each immediately followed by an 8 s pulse to −50 mV to record the tail current. Tail currents were normalized to the maximal block produced by BeKm-1 (ref. 20).

### Optical mapping

On the day of the experiment, the culture medium of cardioids at day 14.5–16.5 was replaced with CardMaintenance medium. To record action potentials, cardioids were incubated for 2 h at 37 °C in preheated CardMaintenance containing 1/100 PowerLoad and 1/1000 FluoVolt membrane potential dye (Invitrogen, #F10488), then transferred to micro-insert wells (IBIDI, #80406) containing 10 µL of preheated CardMaintenance prior to acquisition. To study calcium transients, cardioids were incubated for 15 min at 37 °C in preheated dPBS containing 2 µL mL−1 Pluronic F-127 and 2 µL mL−1 Rhod-2 AM (Invitrogen, #R1244); after three successive 5-min washes in preheated dPBS, cardioids were transferred to micro-inserts containing 10 µL of preheated CardMaintenance and incubated for at least 15 min at 37°C before imaging.

Recordings were performed on an OMS-PCIE-2002 optical mapping system (MappingLab) equipped with two CMOS cameras detecting the action-potential and calcium signals respectively. Acquisitions were performed with OMapRecord 5 software (MappingLab) over a 100 × 100-pixel area corresponding to a maximum surface of approximately 9 mm2 (pixel size 30 µm) at 300 frames per second. For each cardioid, six 20-second sequences were recorded under control conditions; AR-788 was then diluted in 1× CardMaintenance medium maintained at 37 °C, added, and six further 20-second acquisitions were performed at one recording per minute. Raw data were analysed with OMapScope 5 (MappingLab). To improve signal-to-noise ratio, signals were processed by successively applying a Gaussian filter to homogenize fluorescence intensities between neighbouring pixels, a detrending filter to smooth the baseline, and a median filter to reduce electrical noise. Activation maps were generated from the activation time calculated for each pixel, defined as the maximum of the rising slope of the fluorescent signal; conduction velocity was derived from the activation time as an average of three waves. Action-potential duration (APD90), calcium-transient duration (CaD80), interbeat interval and signal amplitude were quantified from the same recordings.

### Compound

The AR-788 (AR) used was N-(1H-benzimidazol-2-yl)-2-(benzylamino)-1,3-thiazole-4-carboxamide (CAS 1232771-02-6; C18H15N5OS; MW 349.41), purchased as a dry powder from ChemDiv (San Diego, CA), catalogue number Y042-4659. ChemDiv guarantees a minimum purity of 90% for catalogue compounds, confirmed by 1H NMR and/or LC(UV)/MS. It was supplied in a labelled glass vial and shipped directly to l’institut du thorax, Nantes. The compound was dissolved in DMSO to obtain a 10 mM stock solution and diluted in the external solution to the desired final concentration; 10 µM was the concentration primarily used, with 5 and 20 µM used for the concentration–response series. Solutions were formulated to maintain an identical final DMSO concentration in both control and treated conditions (≤ 0.1% v/v).

### Statistical analysis

All statistical analyses were performed in GraphPad Prism, and quantitative data are expressed as mean ± standard error of the mean (SEM). The normality of each distribution was verified beforehand using a Shapiro–Wilk test, which determined the choice of subsequent test. For comparison of two independent groups, a Student’s t-test was used when its conditions were met, or a non-parametric Mann–Whitney test when the distribution was not normal. Comparisons involving more than two groups used one-way ANOVA with Tukey’s post-hoc test, or two-way ANOVA with Šidák’s post-hoc test when the interaction between two variables was evaluated. A p-value below 0.05 was considered statistically significant. Concentration–response data were fitted to a Hill equation with maximal block constrained to unity; a three-parameter fit with maximal block free was degenerate on the available points and was not used. Significance is indicated as *p < 0.05, **p < 0.01, ***p < 0.001, ****p < 0.0001.

## Declarations

### Data availability

Source data for every figure panel and table, comprising the individual cell-level and cardioid-level values, are provided with this paper as a Source Data file. All other data are available from the corresponding authors on reasonable request.

### Author contributions

M.A. conceived the study, provided the compound and its rationale, and interpreted the data. Y.G. performed electrophysiology and optical mapping and analysed data. S.G. performed part of the experiments and analysis and reviewed the manuscript. G.L. and N.G. supervised the experimental programme and contributed to design and interpretation. All authors reviewed the manuscript.

