## Supplementary Information for "The ARumenamide AR-788 suppresses the transient outward current in human iPSC-derived right-ventricular Brugada cardioids"

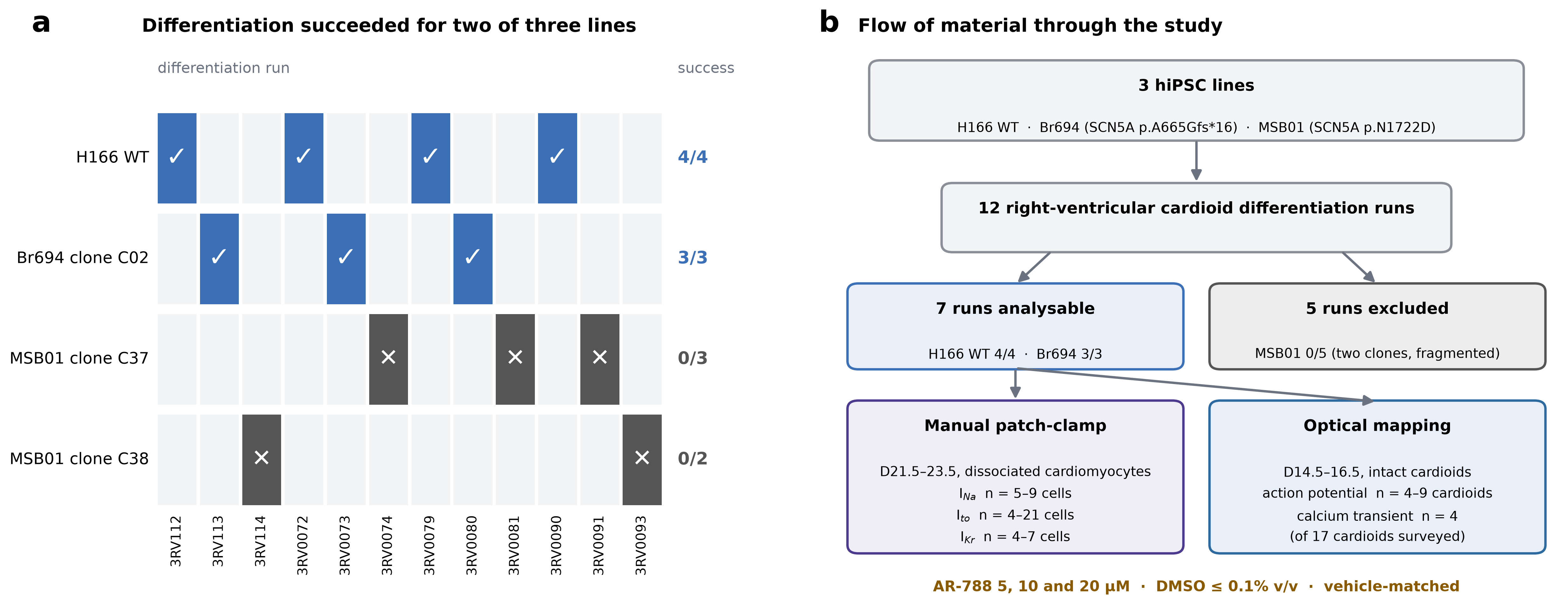

**Supplementary Figure 1. Differentiation efficiency and flow of material through the study.** (a) Every right-ventricular cardioid differentiation attempted in this project, by line and clone. Green, cardioids grew and beat and entered analysis; red, cardioids fragmented and stopped beating. Both independent MSB01 clones failed, so the *SCN5A* p.N1722D missense line contributes no data. (b) Flow of material and the n available to each read-out.

**Supplementary Tables**

**Supplementary Table 1. Baseline electrophysiological phenotype of BrS versus WT cardioids (single-cell patch-clamp and optical mapping).**

| **Parameter** | **WT, mean ± SEM** | **n** | **BrS, mean ± SEM** | **n** | **Δ** | **P** |
| --- | --- | --- | --- | --- | --- | --- |
| Peak INa density at −20 mV (pA pF−1) | −100.6 ± 18 | 9 | −40.67 ± 14 | 7 | −60% | 0.0259 |
| INa slope factor, k | 4.754 ± 0.54 | 5 | 6.409 ± 0.34 | 5 | +35% | 0.031 |
| INa V1/2 activation (mV) | −31.25 ± 2.6 | 5 | −26.46 ± 2.0 | 5 | +4.8 mV | 0.179 |
| Peak Ito density at +60 mV (pA pF−1) | 20.40 ± 2.2 | 21 | 46.38 ± 6.5 | 6 | +127% | <0.0001 |
| Ito / \|INa\| (cohort index) | 0.203 ± 0.042 | n/a | 1.140 ± 0.43 | n/a | 5.6× | n/a |
| Conduction velocity (mm ms−1) | 0.03949 ± 0.0044 | 9 | 0.02458 ± 0.0037 | 8 | −38% | 0.0216 |
| Spontaneous cycle length (ms) | 3734 ± 451 | 8 | 6571 ± 762 | 9 | +76% | 0.0073 |
| Action-potential amplitude (a.u.) | 45.66 ± 8.2 | 9 | 45.10 ± 6.5 | 9 | −1% | 0.958 |
| APD90 (ms) | 343.5 ± 76 | 9 | 332.7 ± 21 | 9 | −3% | 0.892 |
| SD of APD90 (ms) | 25.11 ± 4.1 | 9 | 33.77 ± 2.9 | 9 | +34% | 0.102 |

*Values are mean ± SEM; n = cells (currents) or cardioids (optical mapping). Δ, change relative to WT. The Ito/|INa| index is derived from the group means of two separate cell populations and is not a paired measurement. SD, standard deviation; APD90, action-potential duration at 90% repolarization; a.u., arbitrary units.*

**Supplementary Table 2. Effect of AR-788 (10 µM) on ionic currents in WT and BrS cardiomyocytes.**

| **Parameter** | **Genotype** | **Basal, mean ± SEM** | **AR 10 µM, mean ± SEM** | **Δ** | **n** | **P** |
| --- | --- | --- | --- | --- | --- | --- |
| Peak INa at −20 mV (pA pF−1) | WT | −37.49 ± 3.4 | −33.34 ± 2.8 | −11% | 6 | 0.2101 |
| Peak INa at −20 mV (pA pF−1) | BrS | −15.74 ± 5.1 | −16.39 ± 5.1 | +4% | 6 | 0.3703 |
| Peak Ito at +60 mV (pA pF−1) | WT | 21.48 ± 3.9 | 13.80 ± 3.6 | −35% | 9 | 0.0005 |
| Peak Ito at +60 mV (pA pF−1) | BrS | 42.53 ± 7.4 | 21.71 ± 2.7 | −49% | 4 | 0.0218 |
| Ito, BrS + AR vs WT basal (pA pF−1) | BrS vs WT | 21.48 ± 3.9 | 21.71 ± 2.7 | +1% | 9 / 4 | 0.9716 |
| IKr peak tail current (pA pF−1) | WT | 2.85 ± 0.31 | 1.70 ± 0.29 | −40% | 7 | 0.0035 |
| IKr V1/2 activation (mV) | WT | −33.75 ± 0.97 | −32.65 ± 1.6 | +1.1 mV | 4 | 0.580 |
| IKr slope factor, k | WT | 3.675 ± 0.42 | 4.675 ± 0.62 | +27% | 4 | 0.332 |

*Values are mean ± SEM; n = cells. IKr tail currents are normalized to the maximal BeKm-1–sensitive block; the residual current in BeKm-1 was 1.02 pA pF−1. Absolute INa densities in this table are not comparable with Supplementary Table 1 because the external Na+ concentration was 10 mM here and 20 mM in the phenotyping experiments (see Methods). Row 5 is an unpaired between-genotype comparison of WT basal with BrS + AR, not a before/after test. Basal values are those of the cells that completed the AR protocol (BrS n = 4, WT n = 9) and therefore differ from the phenotyping cohort of Supplementary Table 1 (BrS Ito 46.38, n = 6; WT Ito 20.40, n = 21).*

**Supplementary Table 3. Concentration-dependent inhibition of Ito by AR-788 in WT cardiomyocytes.**

| **AR-788 concentration** | **Ito at +60 mV (pA pF−1), mean ± SEM** | **Normalized Ito (paired), mean ± SEM** | **Inhibition % (paired), mean ± SEM** | **Inhibition % (ratio of means)** | **n** | **P vs basal** |
| --- | --- | --- | --- | --- | --- | --- |
| 0 (basal) | 23.48 ± 5.6 | 1.000 ± 0 | n/a | n/a | 5 | n/a |
| 5 µM | 21.54 ± 6.2 | 0.877 ± 0.046 | 12.3 ± 4.6 | 8.3 | 5 | 0.1274 |
| 10 µM | 17.51 ± 5.5 | 0.703 ± 0.059 | 29.7 ± 5.9 | 25.4 | 5 | 0.0004 |
| 20 µM | 14.58 ± 5.1 | 0.576 ± 0.078 | 42.4 ± 7.8 | 37.9 | 5 | <0.0001 |

*Hill fit constrained to complete block: IC50 = 25.4 ± 3.2 µM, Hill coefficient 1.09 ± 0.17 (fit ± SE); with the Hill coefficient fixed to 1, IC50 = 26.9 ± 1.9 µM. The IC50 lies beyond the highest concentration tested and is an extrapolation. The two inhibition columns are different quantities: the paired column is the mean of each cell's own basal-normalized ratio (the values reported in the text and used for the fit), while the last column is computed from the ratio of the group mean densities in column 2. n = 5 cells carried through all four conditions.*

**Supplementary Table 4. Tissue-level effects of AR-788 (10 µM) by optical mapping of action potentials (AP) and calcium transients (CaT).**

| **Parameter** | **Genotype and modality** | **Basal, mean ± SEM** | **AR 10 µM, mean ± SEM** | **Δ** | **n** | **P** |
| --- | --- | --- | --- | --- | --- | --- |
| Conduction velocity (mm ms−1) | WT, AP | 0.0313 ± 0.0088 | 0.0262 ± 0.0067 | −16% | 4 | 0.2022 |
| Conduction velocity (mm ms−1) | BrS, AP | 0.0164 ± 0.0038 | 0.0185 ± 0.0052 | +13% | 6 | 0.6449 |
| Cycle length (ms) | WT, AP | 8930 ± 3728 | 4679 ± 1026 | −48% | 4 | 0.1041 |
| Cycle length (ms) | BrS, AP | 4293 ± 1607 | 3007 ± 883 | −30% | 6 | 0.1941 |
| APD90 (ms) | WT, AP | 291 ± 131 | 335 ± 139 | +15% | 4 | 0.0996 |
| APD90 (ms) | BrS, AP | 189.3 ± 22 | 195 ± 22 | +3% | 6 | 0.9268 |
| SD of APD90 (ms) | WT, AP | 25.93 ± 6.3 | 32.72 ± 11 | +26% | 4 | 0.4203 |
| SD of APD90 (ms) | BrS, AP | 38.51 ± 8.4 | 28.92 ± 6.0 | −25% | 6 | 0.1142 |
| Conduction velocity (mm ms−1) | BrS, CaT | 0.00880 ± 0.0010 | 0.00562 ± 0.0011 | −36% | 4 | 0.0955 |
| Cycle length (ms) | BrS, CaT | 3385 ± 769 | 3232 ± 1188 | −5% | 4 | 0.8389 |
| Transient amplitude (a.u.) | BrS, CaT | 5.989 ± 0.86 | 5.873 ± 1.0 | −2% | 4 | 0.7066 |
| CaD80 (ms) | BrS, CaT | 640 ± 22 | 629.5 ± 72 | −2% | 4 | 0.8601 |
| SD of CaD80 (ms) | BrS, CaT | 72.84 ± 11 | 63.50 ± 6.9 | −13% | 4 | 0.1469 |

*Values are mean ± SEM; n = cardioids. All comparisons are non-significant. Calcium transients were recorded only in BrS cardioids, so the calcium-transient rows have no WT arm. Panels are exploratory owing to small n (see Methods).*

**Supplementary Table 5. Molecules used for cardioid generation.**

| **Molecule** | **Supplier** | **Reference** |
| --- | --- | --- |
| ROCKi | Tocris | #1254 |
| LY294002 | Tocris | #RD-1130/5 |
| Insulin | Roche | #11376497001 |
| CHIR99021 | R&D Systems | #RD-4423/50 |
| BMP4 | Miltenyi Biotec | #130-111-164 |
| Activin A | Miltenyi Biotec | #130-115-010 |
| FGF2 | Miltenyi Biotec | #130-093-564 |
| SB431542 | Tocris | #1614/10 |
| Retinoic acid (RA) | Sigma-Aldrich | #R2625 |
| C59 | Tocris | #5148/10 |
